# Climatic influences on the worldwide spread of SARS-CoV-2

**DOI:** 10.1101/2020.03.19.20038158

**Authors:** Michail Bariotakis, George Sourvinos, Elias Castanas, Stergios A. Pirintsos

## Abstract

The rapid global spread of the novel, pathogenic, SARS-CoV-2 causing the severe acute respiratory disease COVID-19, becomes a major health problem worldwide and pose the need for international predictive programs. Given the lack of both specific drugs and an efficient preventive vaccine, the expectation that SARS-CoV-2’s transmission rate might decrease in temperate regions during summer, dominated the social scene. Here, we attempted a prediction of the worldwide spread of the infections based on climatic data, expressed by 19 bioclimatic variables. The calculated probability maps shown that potential areas of infection follow a shift from the Tropical to Temperate and Mediterranean Bioclimatic regions, and back to the Tropics again. Maps show an increased probability of infections in Europe, followed by an expansion covering areas of the Middle East and Northern Africa, as well as Eastern coastal areas of North America, South-Eastern coastal areas of Latin America and two areas of Southern Australia, and later return to areas of Southeastern Asia, in a manner similar to that of influenza strains (H3N2). Our approach may therefore be of value for the worldwide spread of SARS-CoV-2, suggesting an optimistic scenario of asynchronous seasonal global outbreaks, like other viral respiratory diseases. Consequently, we suggest the incorporation of a climatic impact in the design and implementation of public health policies. Maps of our model are available (constantly updated up to the saturation of the model) at: https://navaak.shinyapps.io/CVRisk/.

## Introduction

The recent emergence of the novel pathogenic SARS-coronavirus 2 (SARS-CoV-2), causing the severe acute respiratory disease COVID-19 and its rapid intra-national and international spread, as well as its pandemic assessment by WHO, pose a global health emergency^1,2^. The human-to-human transmission, mainly via the respiratory route, the pace of spread, as well as the description of infected individuals suggest that SARS-CoV-2 is quite contagious^3^. Given the current lack of both specific effective drugs and an efficient preventive vaccine, Health Authorities have focused on public health management measures for the restriction of the viral spread.

Based on the seasonal behavior of the SARS-CoV-2’s closest relatives beta-coronaviruses OC43 and HKU1, it is expected that SARS-CoV-2’s transmission rate might decrease in temperate regions during summer and will follow a similar seasonal pattern as the other respiratory viruses. However, newly population-introduced viruses may behave differently, their propagation being dependent on factors such as the environment, the human behavior, the host’s immune system and the depletion of susceptible hosts, making this expectation problematic. The situation is further complicated by the lack of accurate long-term meteorological forecasts.

Meteorological data provide short term (e.g. day, week, and month within a year) variations of a measured finite number of parameters (such as wind, precipitation and temperature, among others) in a given geographical area, and are used for short-term weather variation forecasts. In contrast, climate data are collected over a number of years (usually 30 years) and provide the mean conditions in a given area, over decades. In the present report, we have attempted the prediction of viral spread as a function of these climate parameters.

We built a predictive model of the potential global distribution of SARS-CoV-2, based on data of virus detection/infected individuals worldwide, using data from the World Health Organization (WHO) over time, and bioclimatic data as variables. We fitted an entropy model, integrating all bioclimatic parameters and show that climate may successfully predict (with a sensitivity >90%) the outbreak of SARS-CoV-2 spread, in different geographic areas. Our model supports therefore a climate-related model of SARS-CoV-2 spread. We propose that such a model, continuously updated, might be a useful tool for the establishment of appropriate public health measures.

## Results

We have used the number of infected cases, provided in all WHO Situation Reports, covering the period 02 February to 04 March, 2020 and fitted them in a maximum entropy model with all 19 bioclimatic variables as cofactors (Table 1). The model, reiterating per five days with additional reported data released by WHO, provides the probability of infection, in each geographical point, worldwide. The obtained probability maps are presented in Figure 1A for the period discussed here, while on the accompanying web site, these data are continuously updated every five days, after any new WHO release. As shown, potential areas of infection follow a shift towards Europe, which becomes more pronounced with time. This shift is followed by an expansion, covering areas of the Middle East and Northern Africa, as well as the Eastern coastal areas of North America, the South-Eastern coastal areas of Latin America and two areas of Southern Australia. Interestingly, at the later measures a high probability of novel infections in China regions and in Southwest Asia is also predicted (Figure 1B).

**Table 1.** List of the bioclimatic variables used in the construction of the model.

| Variable | Description | Temporal Scale |
| --- | --- | --- |
| BIO1 | Annual Mean Temperature | Annual |
| BIO2 | Mean Diurnal Range (Mean of monthly (max temp - min temp)) | Variation |
| BIO3 | Isothermality (BIO2/BIO7) (* 100) | Variation |
| BIO4 | Temperature Seasonality (standard deviation *100) | Variation |
| BIO5 | Max Temperature of Warmest Month | Month |
| BIO6 | Min Temperature of Coldest Month | Month |
| BIO7 | Temperature Annual Range (BIO5-BIO6) | Annual |
| BIO8 | Mean Temperature of Wettest Quarter | Quarter |
| BIO9 | Mean Temperature of Driest Quarter | Quarter |
| BIO10 | Mean Temperature of Warmest Quarter | Quarter |
| BIO11 | Mean Temperature of Coldest Quarter | Quarter |
| BIO12 | Annual Precipitation | Annual |
| BIO13 | Precipitation of Wettest Month | Month |
| BIO14 | Precipitation of Driest Month | Month |
| BIO15 | Precipitation Seasonality (Coefficient of Variation) | Variation |
| BIO16 | Precipitation of Wettest Quarter | Quarter |
| BIO17 | Precipitation of Driest Quarter | Quarter |
| BIO18 | Precipitation of Warmest Quarter | Quarter |
| BIO19 | Precipitation of Coldest Quarter | Quarter |

**Figure 1.**
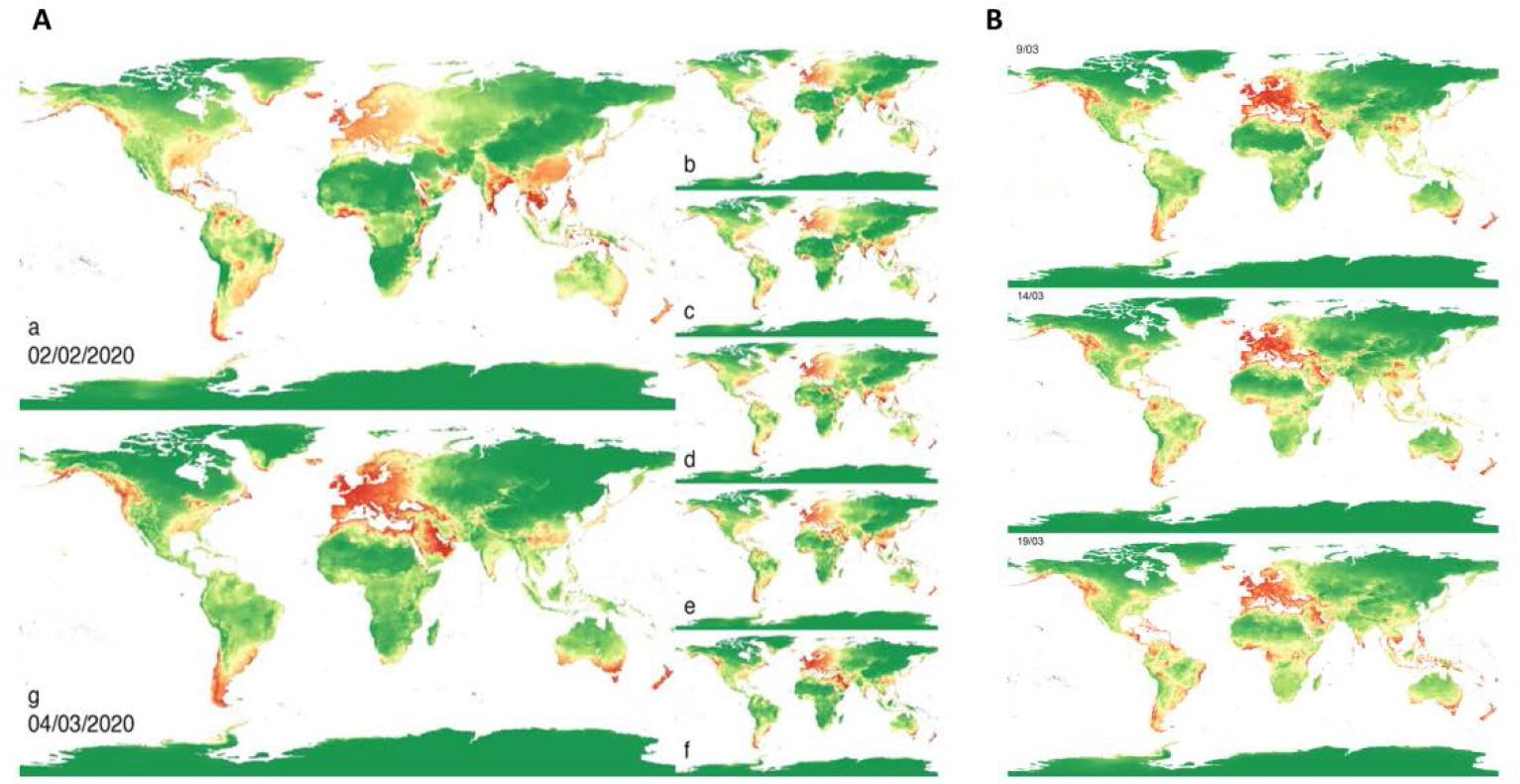
**A**. Potential distribution of SARS-CoV-2 infections. Each map represents predictions of a model trained with data from all countries with at least 1 (total) confirmed case for the respective situation report. Latin letters a-g represent the temporal order (2, 7, 12, 17, 22, 27 February and 4 March respectively). Red corresponds to the highest probability of viral presence, while green corresponds to lowest. **B**. Potential distribution of SARS-CoV-2 infections of the last three intervals (9, 14, 19 March) before the saturation of the model. Red corresponds to the highest probability of viral presence, while green corresponds to lowest.

With the integration of new cases, the precision of the model increases (Figure 2A), attaining a positive predictive value over 90%; the model correctly predicted the outburst of infections in Europe and California. However, it should be noted that the potential predicted infections might not always coincide with the actual ones. Such discrepancies can provide valuable insights into the underlying mechanisms and barriers of infection spread. Additionally, as shown, the prediction of the model attains the value of 1, at latter measures (dates after March 9), suggestive of a model saturation, as infected cases are reported in all countries all over the world.

**Figure 2.**
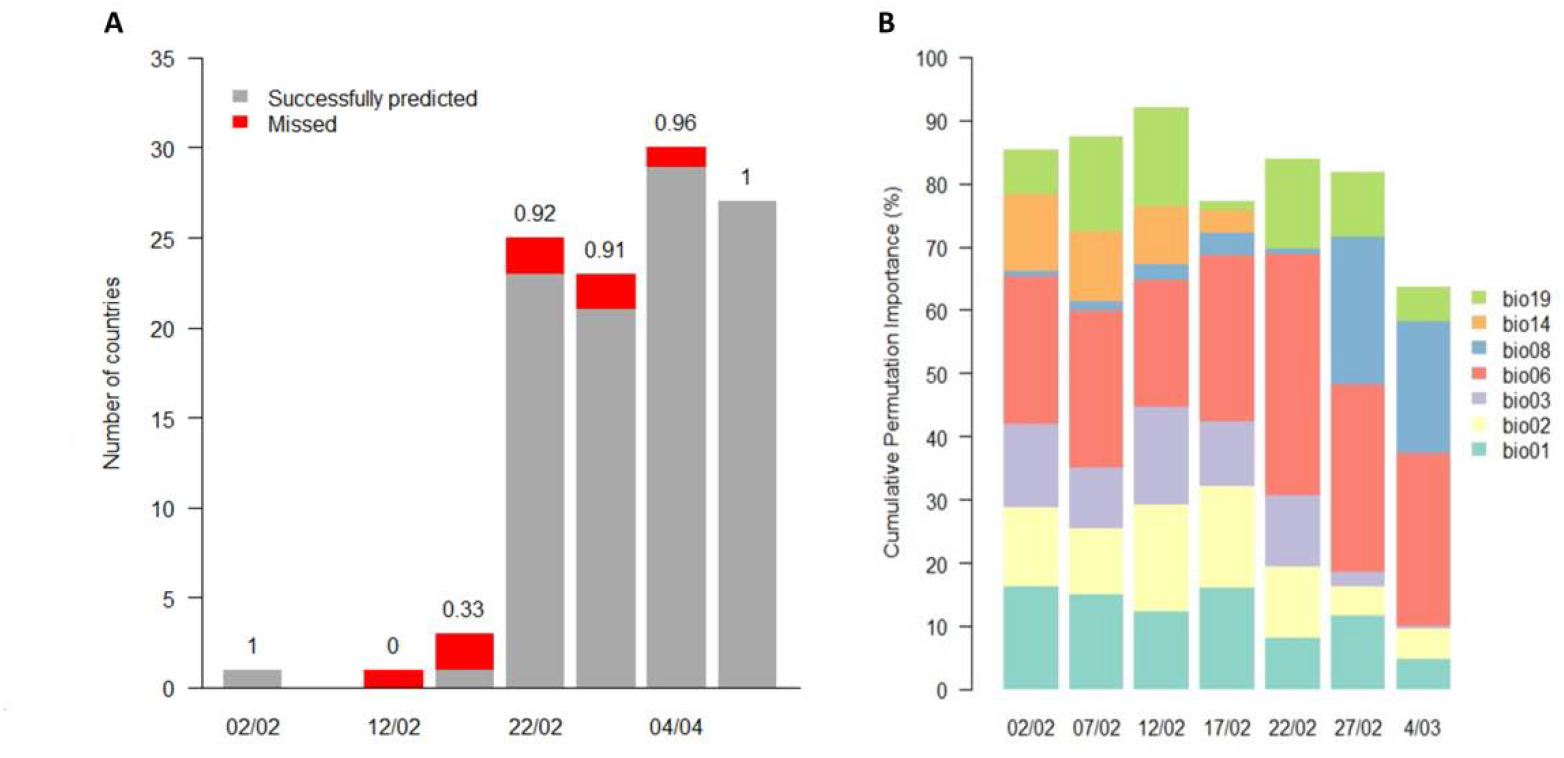
Importance of the most dominant bioclimatic predictors for each model (permutation analysis). Only predictors with mean importance of at least 5% are shown. Variables are as follows: bio1 = annual mean temperature, bio2 = mean diurnal range, bio3 = isothermality, bio6 = min temperature of the coldest month, bio8 = mean temperature of the wettest quarter, bio14 = precipitation of the driest month, bio19 = precipitation of the coldest quarter.

Following the impact of each bioclimatic variable in the model (Figure 2B), we observe that a small subset of variables can predict the viral outbursts, at a given time-point. In addition, there is a notable shift of the predictive power of some parameters. In particular, the parameter “Precipitation of Driest Month” (bio14) is losing importance and is replaced by “Mean Temperature of Wettest Quarter” (bio8). Moreover, the importance of “Isothermality” (bio3), which quantifies how large the day-to-night temperatures oscillate relative to the summer-to-winter (annual) oscillations, is highly reduced during the period of the more recent WHO Situation Reports. It is to note that an isothermal value of 100 indicates the diurnal temperature range is equivalent to the annual temperature range, while anything less than 100 indicates a smaller level of temperature variability within an average month relative to the year. The two major predictive variables, at the current state of viral spread (data of March 4) are the bio6 (min temperature of the coldest month, permutation importance 27.4%) and bio8 (mean temperature of the wettest quarter, permutation importance 20.9%).

## Discussion

Data presented here, show that SARS-CoV-2 distribution/reported infections can be efficiently predicted by a small subset of bioclimate environmental parameters. Our model, however, is based on data released by WHO at a country level. Its predictive power could increase with the incorporation of data with a higher spatial precision (eg. at the regional or city level). An analogous methodology has been followed in a preprint manuscript which appeared in medRxiv^4^.

An integration of world Bioclimates of each continent was recently reported^3^. Under this concept, we predict a shifting from Tropical to Temperate and Mediterranean bioclimatic regions, which corresponds to a transition from low (in Tropical Bioclimate) to high yearly seasonality (especially in Mediterranean Bioclimate). This could be a driving force of virus propagation. In addition, a scale shift to Boreal Bioclimates is recorded, especially in the areas of North-Eastern coastal areas of North America. It is not clear whether this is a marginal prediction of our model but it is worthy of mention that the burden of epidemics in these areas, taking into consideration the epidemic events globally for the period 2011-2017, are the highest worldwide^5^.

The shifting of the spread at a global scale pose the need for further investigation towards the identification of the underlying relationships of the bioclimatic variables with the virus-host interactions. Nevertheless, our results so far contribute to further understanding the viral impact worldwide. Specifically, it becomes clear from the very early predictive maps that the virus doesn’t restrict to the tropical and nearby areas as the Ebola virus does, as evidenced by its rather limited penetration in tropical Africa and South America regions. Starting from the city of Wuhan, in Hubei province, it spreads worldwide, following a scheme that has been suggested for seasonal viruses^6^.

A common model in viral ecology is that of sink–source model, where a source is located in the tropics and sinks in temperate regions^6,7^. This model designates that Southeastern Asia may function as a reservoir that maintains high diversity of influenza viruses that seed seasonally to other temperate regions. If this scheme accurately describes the overall pattern of SARS-CoV-2 spread, then its antigenic characteristics should be investigated within Southeastern Asia, in view of the development of an efficient vaccine. However, in an alternative interpretation of our data, there might not exist any restrictions of viral migration between temperate and tropical regions, suggesting a continuous bidirectional spread between tropic and temperate regions, as indicated by our prediction (verified by recent data) of a secondary spread to areas of Southeastern Asia and subtropical regions of China. Such a distribution is not in complete agreement with a sink–source model of viral ecology. A similar viral distribution has indeed proposed for influenza A H3N2. It has been suggested that the global influenza population is maintained by a series of temporally overlapping epidemics, coupled with high rates of migration, supported by an unevenly distributed network of flights, connecting highly dense urban centers^8,9^. This scheme suggests a more complicated spread of viral antigenic variants, which present an obstacle for the improvement of vaccine strain selection. In this scheme, a second coronavirus wave in Hubei province is predicted, and currently confirmed. It might, therefore, be interesting to follow whether the virus might have difficulty reestablishing itself in the community, taking into consideration the community immunity impact, generated by the primary infection.

In both schemes, asynchronous seasonal global outbreaks are expected, like in other respiratory diseases. However, it is worthy of mention that this coronavirus is not a biological or ecological singularity, as it seems to follow similar global circulation patterns with other influenza viruses, an element which might be of value in designing global health policies.

The simulations presented in Figure 1 are available and gradually upgraded at the following link: https://navaak.shinyapps.io/CVRisk/.

## Methods

### The maximum-entropy approach

In total, we used data from seven WHO Coronavirus Disease-2019 Reports (Situation Report −8, −13, −18, −23, −28, −33, −39, and −44), which spanned the period from 02/02/2020 to 04/03/2020 on a regular interval of five days. Subsequent maps of the next three intervals before the saturation of the model were also produced based on the Situation Report −49, − 54, and −59. The countries with at least one confirmed incident in each Situation Report or any of the previous ones were used as “presence records” of the virus.

We also employed the widely used bioclimatic data from WorldClim.org^10^ in 10 minutes (corresponding to approximately 18.5 km) resolution for the whole world map. WorldClim is a set of global climate layers (gridded climate data), which can be used for mapping and spatial modeling. These data represent variables derived from climatic data, which are considered to have an important effect on biological entities (see Table 1).

To correlate the virus presence records given by WHO with the bioclimatic variables, we applied a machine-learning technique called maximum entropy modeling, employing Maxent^11^ version 3.4.1. Maxent was initially designed to model the potential niche of species, given a set of presence records and a set of environmental predictors. The model expresses a probability distribution where each grid cell has a predicted suitability of conditions for the species, with the underlying assumption that the species relates to the employed variables, in our case to the bioclimatic variables.

Analysis was performed in R version 3.6.0^12^, which was also used for the creation of the Figures and Maps.

### Data availability

Concerning SARS-CoV-2, data are available from the official site of WHO: https://www.who.int/emergencies/diseases/novel-coronavirus-2019/situation-reports

Bioclimatic data are available from https://www.worldclim.org/

## Data Availability

Data are online available from the sources which are described in the text

## Notes

### Competing Interest Statement

The authors have declared no competing interest.

### Funding Statement

No funding support

